# Obtaining Actigraphy Sleep Report Using Garmin Wearable Devices and Labfront App

**DOI:** 10.1101/2023.11.03.23297889

**Authors:** Chang Francis Hsu, Han-Ping Huang, Chris Peng, Jordan Masys, Andrew C. Ahn

**Author notes:** **Corresponding Author:** Andrew C. Ahn, Labfront (Kiipo Co.), Boston, MA, 02067, USA.

## Abstract

In a collaborative effort between Labfront and Garmin, an actigraphy-based sleep analysis report has been developed for Garmin’s consumer-grade wearable devices, accessible via the Labfront application. Comparative analyses against established medical-grade actigraphic devices, namely Actiwatch-2 and Motionlogger, demonstrate notable concordance in key performance indicators. Specifically, the Garmin device yielded a sensitivity of over 95% for sleep detection, a specificity of over 80% for wake detection, and an overall accuracy exceeding 95%.

The study introduces a transparent methodology for deriving sleep metrics from acceleration data, thereby addressing a current gap in the literature regarding consumer-grade actigraphy solutions. Additionally, the system allows for individual customization of sleep/wake detection parameters, offering adaptability to specific user conditions. These findings suggest that consumer-grade wearable devices may serve as viable alternatives to medical-grade actigraphs for sleep analysis, without sacrificing analytical rigor.

## 1. Introduction

Actigraphy serves as a valuable tool for monitoring sleep patterns by recording wrist movement through accelerometers. In clinical settings, devices like the Actiwatch-2 (Phillips-Respironics, Bend, Oregon) and the Motionlogger Sleep Watch (AMI, NY) have become the gold standard. An overwhelming 86% of actigraphy research between 2012 and 2020 utilized these medical-grade devices, according to a review by Schoch [1]. However, their high cost and lack of transparent methodology pose challenges for both clinicians and general users. The situation has been further complicated by Philips’ recent announcement in October 2002 to “discontinue offering wrist-worn products in the clinician and research markets”, making all Actiware, Actiwatch-2, Actiwatch PRO, and Actiwatch Plus products unavailable for purchase after Dec. 29, 2022.

Addressing this gap, Labfront and Garmin embarked on a series of studies to assess whether consumer-grade wearables, like the Vivosmart-4 (Garmin, Olathe, Kansas) could effectively substitute these medical-grade actigraphs [2]. Steps were made to deduce the algorithm to the raw-ACC data obtained from Vivosmart-4. Our findings suggest promising agreement between the consumer-wearable and medical-grade actigraphs, opening doors for more affordable and accessible sleep monitoring using commercial wearable devices with the Labfront app.

In this manuscript, we will first delve into the fundamentals of actigraphy. We will then detail how we converted raw acceleration data into sleep quality metrics using the Garmin Vivosmart-4, comparing its performance with the Actiwatch-2 and Motionlogger. Lastly, we will present a sample actigraphy sleep report generated by this approach.

## 2. Background and Method

### 2.1. Introduction to Actigraphy

Actigraphy is a non-intrusive method for capturing limb movement over extended periods, typically ranging from days to weeks. While actigraphic devices can be affixed to various parts of the body such as the wrist, ankle or waist, the wrist is the typical location for sleep assessment. Mathematical algorithms are then applied to the recorded acceleration data to estimate periods of wakefulness and sleep. Actigraphy offers two major advantages over in-lab polysomnography (PSG), the gold standard in sleep measurement: 1) It allows for the observation of sleep behavior in a natural home setting, and 2) It enables continuous, 24-hour monitoring over longer time spans.

Actigraphy is particularly useful in diagnosing and monitoring a range of sleep-related disorders, such as insomnia, sleep apnea, sleep movement disorder, and circadian rhythm disturbances. For a more comprehensive introduction to actigraphy, you may refer to our previous post, Introduction to Actigraphy [3].

### 2.2. Quantifying Activity from Acceleration Signal

Two prevalent approaches for quantifying activity are Proportional Integrating Mode (PIM) and Zero Crossing Mode (ZCM).

PIM operates by sampling the accelerometer’s output signal at a high frequency and then computing the area under the curve [4-5]. Figures 1a and 1b illustrate the acceleration signal of periodic hand-shaking with different amplitudes. There are 10 hand-shakes over a 5-second span with an amplitude of approximately 200 (Figure 1a) and 100 mG (Figure 1b). The cumulative area under the curves is 600 and 350 mG • s, respectively. PIM is generally regarded as a measure of activity magnitude or motion vigor.

**Figure 1.**
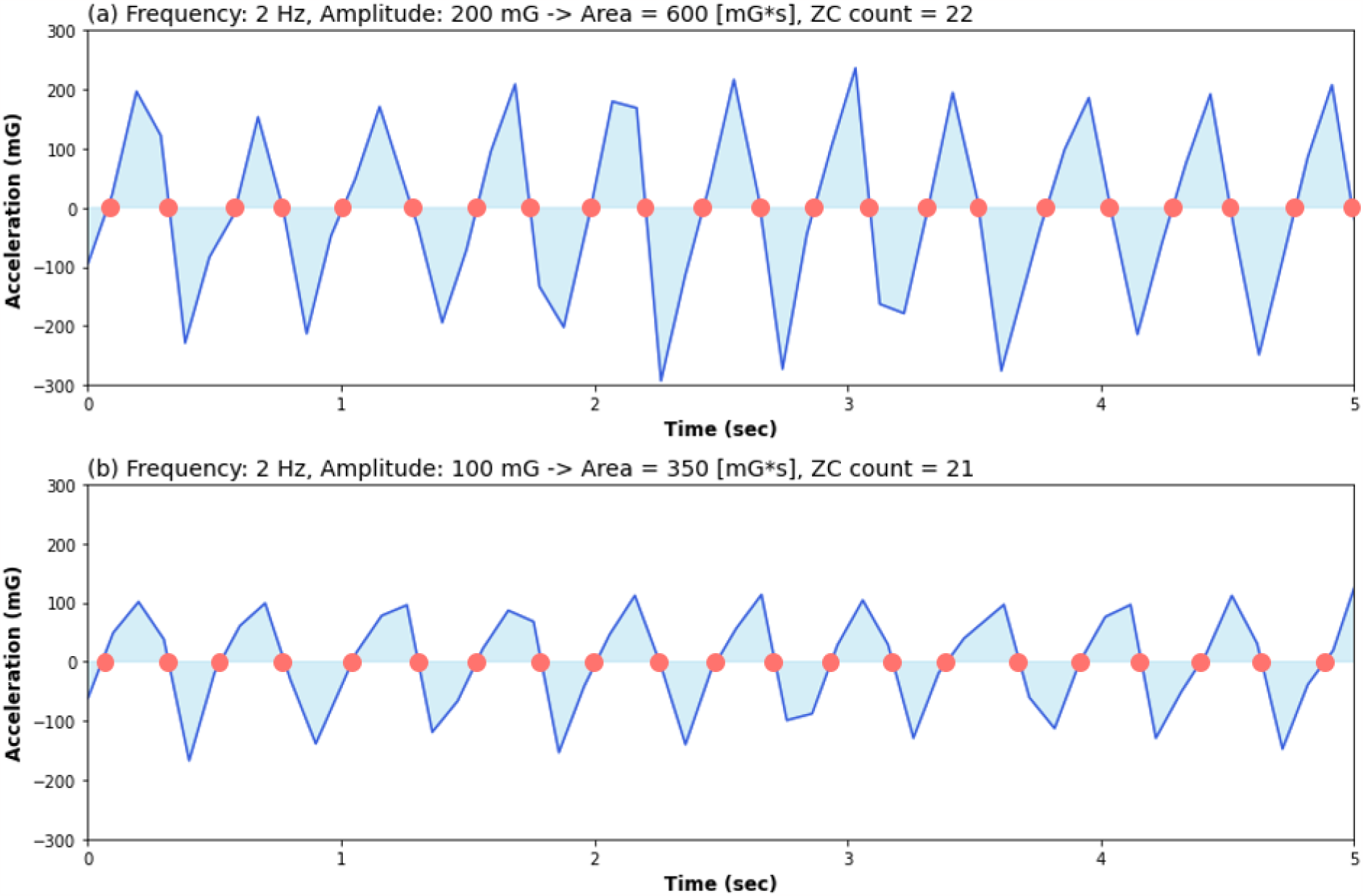
Acceleration signal of periodic handshaking. The blue regions indicated the area under the curve, and the red circles indicated the signal crossed zero.

Conversely, Zero Crossing Mode (ZCM) counts the number of times the signal crosses zero (or near zero), as marked by red circles in Figure 1. Notably, even though Figure 1a has a higher amplitude than Figure 1b, both exhibit similar frequencies of movements and ZC counts. Hence, while PIM quantifies the amplitude of motion, ZCM focuses on its frequency.

### 2.3. Actigraphic Devices

Motionlogger Sleep Watch (AMI, NY) and Actiwatch-2 (Phillips-Respironics, Bend, Oregon) are the two most commonly used medical-grade actigraphic devices for sleep analysis [1,4,6]. While Motionlogger employs both Zero Crossing Mode (ZCM) and Proportional Integrating Mode (PIM), Actiwatch-2 relies exclusively on PIM. Motionlogger’s User Guide recommends ZCM for its high accuracy [7]. As previously noted, Actiwatch-2 became commercially unavailable when Philips discontinued its entire line of wrist-worn products after December 29, 2022.

Generally, these medical-grade actigraphs are financially out of reach for the average consumer, each carrying a price tag of approximately $3,000, which covers both hardware and software costs (as of the time this manuscript was submitted). The lack of transparency regarding the chosen sleep parameters - used to derive sleep measures - adds a layer of complexity for clinicians and researchers. Nonetheless, these devices do provide some level of adjustability, allowing users to modify settings such as sampling rate, epoch length, and sensitivity, often categorized into low, medium, or high thresholds.

Priced at $150 USD (as of this manuscript’s submission), the Vivosmart-4 by Garmin presents a cost-effective alternative to medical-grade actigraphs. Users of the Labfront app can now benefit from customizable settings essential for accurate actigraphic sleep analysis. These include options for epoch window size, IIR filtering techniques, methods for calculating acceleration vector magnitude, and the choice of quantification approach (zero-crossing mode, proportional integration mode, or time above thresholds), as well as threshold sensitivity levels. Furthermore, the device enables the concurrent collection of three separate time-series, each with its own unique settings, thereby simplifying the process of identifying the most effective, personalized configurations for users.

With the introduction of these customizable features, this study aims to assess whether this consumer-grade alternative can yield results comparable to those obtained with Actiwatch-2 and Motionlogger in a study participant. In Sections 3 and 4, we will detail the methodology for deriving sleep quality parameters from acceleration data using the Garmin wearable device, followed by a comparison of results with those from Actiwatch-2 and Motionlogger. We will conclude by presenting a sample actigraphy sleep report.

### 2.4. Methodology of Sleep/Wake Detection Using PIM and ZCM

The flow charts of deriving sleep/wake detection using PIM and ZCM modes are similar, as shown in Figure 2. We will describe the process of generating the Sleep-Wake detection step-by-step accordingly.

**Figure 2.**
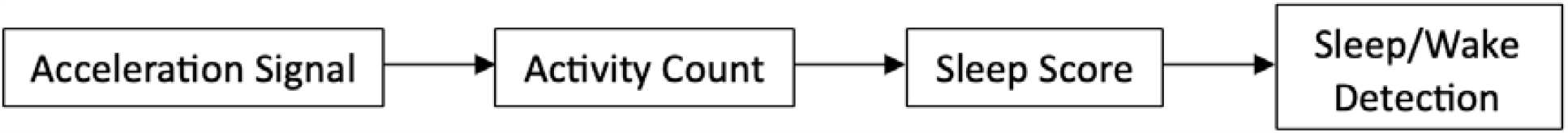
The flow charts of deriving sleep/wake detection.

#### 2.4.1. Proportional Integration Mode (PIM)

Figure 3 outlines the signal processing method used for determining sleep/wake patterns, further detailed in the referenced literature [8-10]. Figure 3(a) displays the 3-axis acceleration signal from the Vivosmart-5, sampled at 25Hz and filtered through a 3-11 Hz bandpass. Figure 3(b) shows the time series of activity counts, denoted as A_P_(n), where ‘n’ represents the epoch index with a 30-second epoch length. The calculation of A_P_(n) involves two steps. First, acceleration values falling below a user-adjustable noise threshold are set to zero. Subsequently, the acceleration values within each 30-second epoch are summed to quantify the activity count for that epoch.

**Figure 3.**
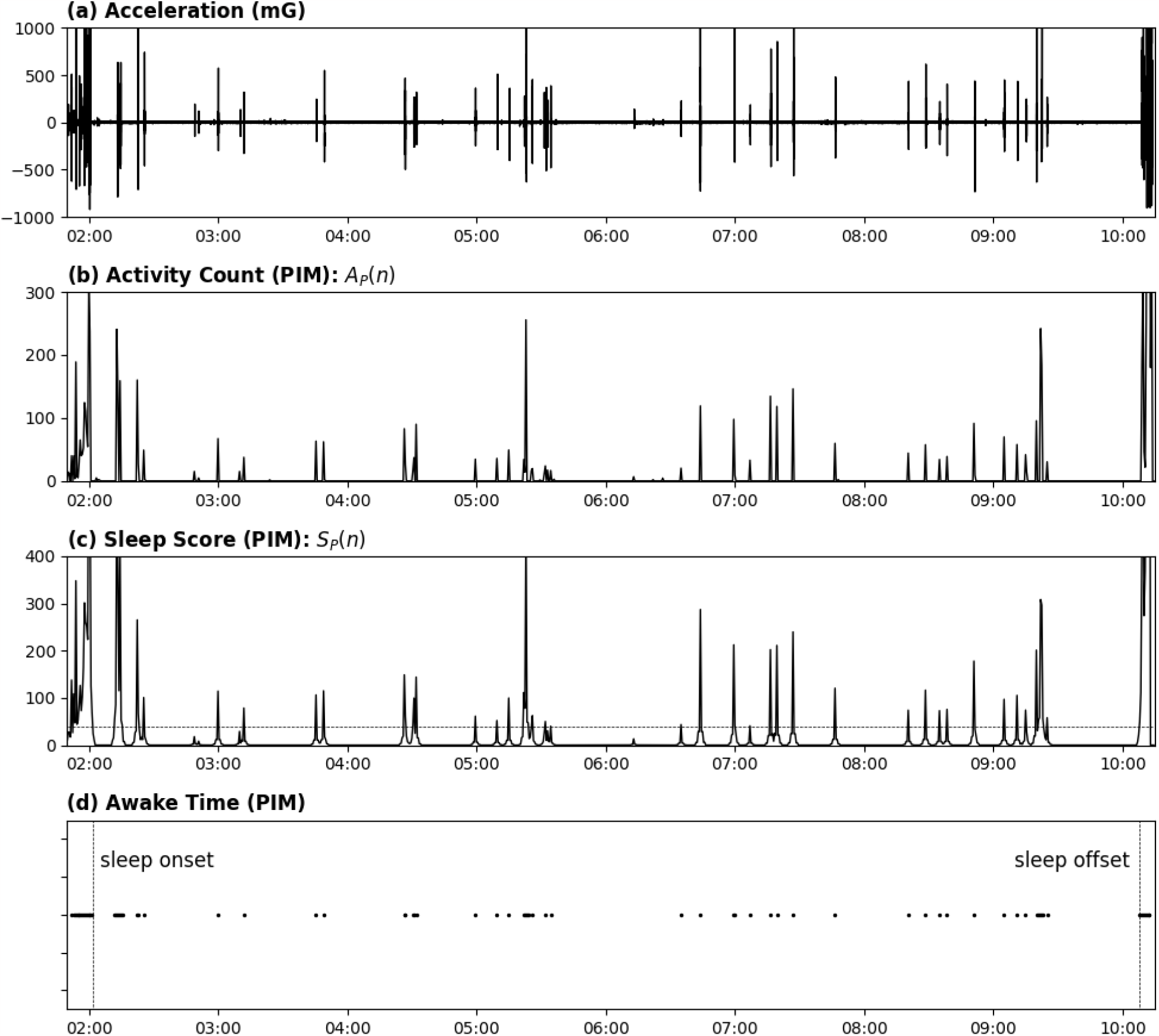
Acceleration Signal to Sleep/Wake Detection (PIM).

Figure 3(c) illustrates the sleep score labeled as S_P_(n), which is derived from A_P_(n) [10]:

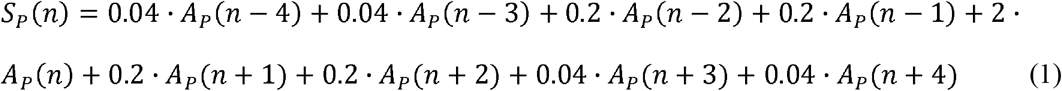

Ultimately, epochs with S_P_(n) values below 40 are identified as sleep periods, while those above 40 are classified as wake periods. The threshold value of 40 is the default setting for sleep/wake delineation as established in reference [10]. Figure 3(d) accordingly displays the resulting sleep/wake patterns.

#### 2.4.2. Zero-Crossing Mode (ZCM)

Figure 3 depicts the signal processing technique used to derive sleep/wake patterns using the ZCM mode. The underlying methodology is detailed in reference [4,7].

Figure 4(a) displays the same acceleration signal as presented in Figure 3(a). Figure 4(b) shows the activity count per minute, designated as A_Z_(n), where ‘n’ represents the epoch index with a one-minute epoch length. In Zero Crossing Mode (ZCM), the activity count corresponds to the number of times the signal crosses or closely approaches zero (or near zero)

**Figure 4.**
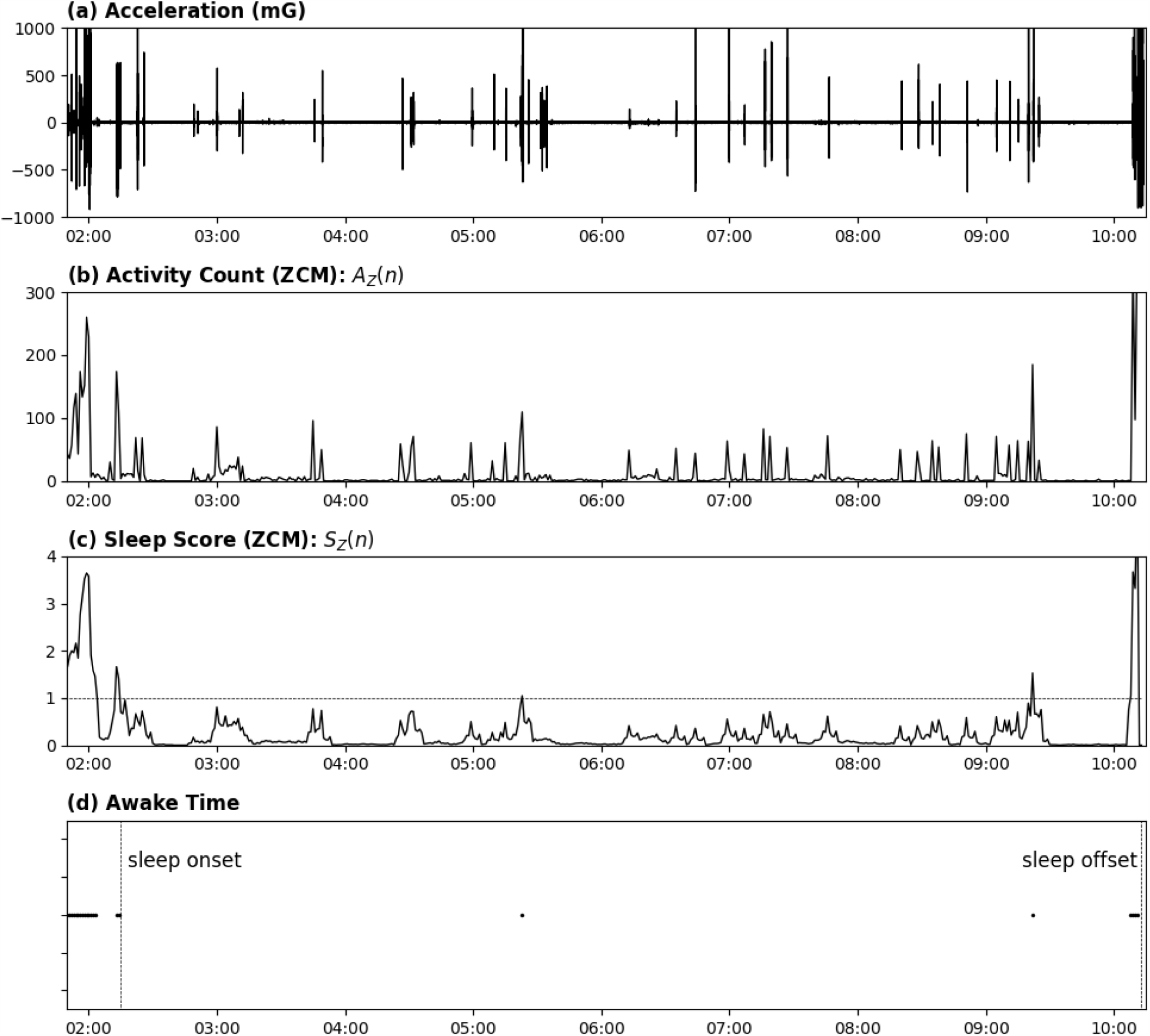
Acceleration Signal to Sleep/Wake Detection (ZCM).

Figure 4(c) illustrates the sleep score labeled S_P_(n), which is derived from A_Z_(n) [4,7]:

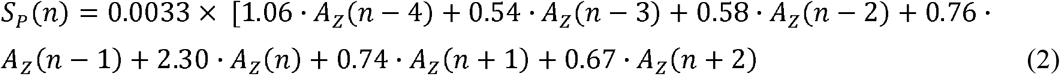

In the end, epochs with S_P_(n) values below and above 1 are identified as sleep and wake periods, respectively. Figure 4(d) depicts the derived sleep/wake patterns.

Table 1 presents the sleep parameters extracted using PIM (as shown in Figure 3) and ZCM (as depicted in Figure 4). Notably, there are significant differences in Wake After Sleep Onset (WASO) and Wake Episodes (WE). Further comparisons will be elaborated upon in Section 3.

**Table 1.** Sleep Parameters and Comparison between PIM and ZCM.

|  | Description [4,10] | Unit | PIM | ZCM |
| --- | --- | --- | --- | --- |
| **Bed-Time** | From Sleep survey | Time | 1:30 |  |
| **Get-Up** | From Sleep survey | Time | 10:14 |  |
| **Sleep Onset** | PIM: The time point of the start of the first 10 consecutive minutes of sleep after getting to bed [10]. | Time | 2:02 | 2:15 |
|  | ZCM: The time point of the first continuous block of at least 20 min of sleep with no more than 1 min of interruption [4] |  |  |  |
| **Sleep Offset** | PIM: The time point of the last minute of the last 10 consecutive minutes of sleep before getting out of bed [10]. | Time | 10:08 | 10:13 |
|  | ZCM: The last minute the subject was scored asleep before getting out of bed [4]. |  |  |  |
| **Time in Bed** | (Get Up Time) - (Bed Time) | min | 525 |  |
| **O-O interval** | (Sleep Offset) - (Sleep Onset) | min | 486 | 478 |
| **Wake After Sleep Onset (WASO)** | # of minutes of wake during O-O interval | min | 28 | 6 |
| **Total Sleep Time** | (O-O interval) - WASO | min | 458 | 472 |
| **Sleep Onset Latency** | (Sleep Onset) - (Bed Time) | min | 32 | 45 |
| **Sleep Efficiency** | (Total Sleep Time) / (O-O Interval) | % | 0.94 | 0.99 |
| **Wake Episodes** | # of switching from sleep to wake | # | 37 | 3 |

## 3. Result

### 3.1. Comparison among Motionlogger, Actiwtch-2, and Vivosmart-5

The configuration of our experiment is depicted in Figure 5, where Motionlogger, Actiwatch-2, and Vivosmart-5 were worn concurrently during sleep for a comparative analysis. The Motionlogger and Actiwatch-2 employ piezoelectric accelerometers with sampling rates of 10 Hz and 32 Hz, respectively. In contrast, the Vivosmart-5 utilizes a 3-axis MEMS accelerometer with a sampling rate of 25 Hz.

**Figure 5.**
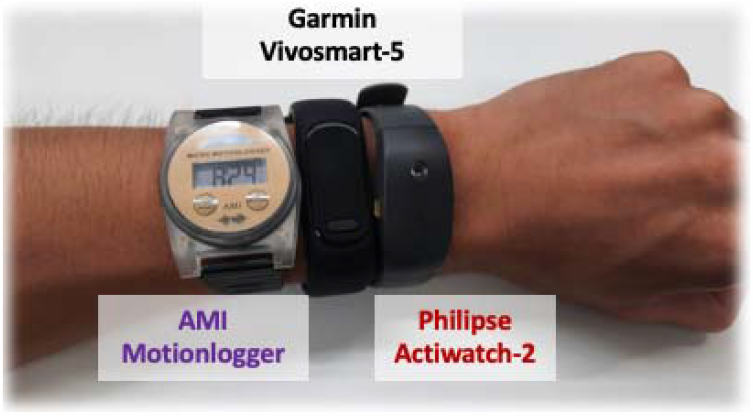
Motionlogger, Actiwatch-2, and Vivosmart-5 on the participant’s right wrist during sleep.

Figure 6 illustrates the sleep/wake patterns of the 3 nights derived from Actiwatch/Motionlogger and Garmin wearable.

**Figure 6.**
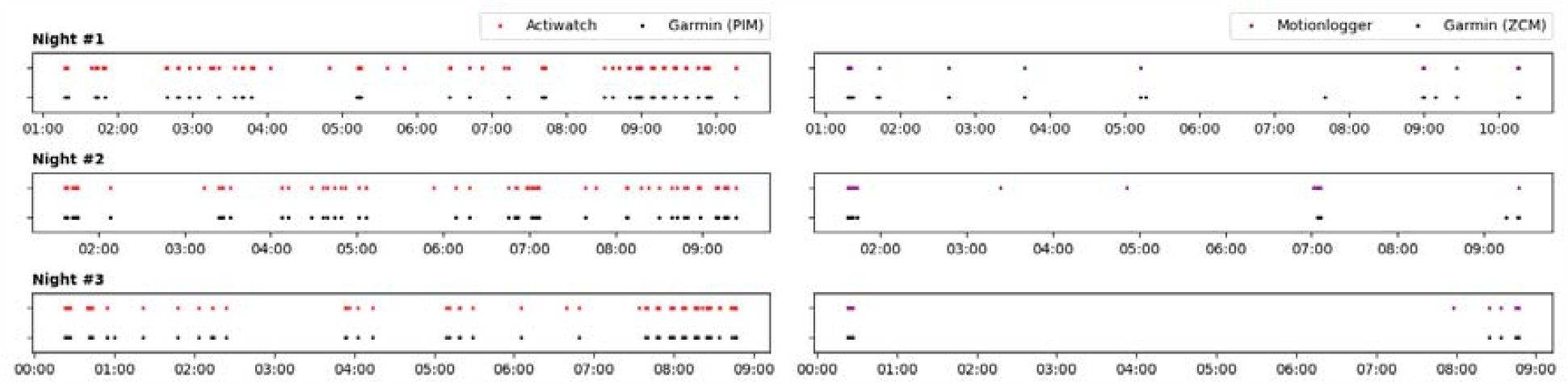
Sleep/Wake Patterns of 3 nights derived from 3 wearable devices. The labels indicate the awake time.

Table 2 presents the consistent and inconsistent outcomes across the three wearable devices under study. The Garmin Vivosmart-5 shows a high degree of agreement with both Actiwatch-2 and Motionlogger, revealing an accuracy of 0.98 and a specificity greater than 0.8 for wake detection. However, the consistency between Actiwatch-2 and Motionlogger is notably lower, with a specificity measuring just 0.32.

**Table 2.** Confusion matrices of inter-device comparison in sleep/wake detection.

| (a) |  | Actiwatch<br>(Treating as standard) |  |
| --- | --- | --- | --- |
|  |  | Sleep (min) | Wake (min) |
| **Vivo.** | S | 1386 (TP) | 22 (FP) |
| **(PIM)** | W | 9 (FN) | 89 (TN) |

|  |  | Motionlogger |  |
| --- | --- | --- | --- |
|  |  | Sleep | Wake |
| **Vivo.** | S | 1465 | 7 |
| **(ZCM)** | W | 7 | 33 |

|  |  | Actiwatch |  |
| --- | --- | --- | --- |
|  |  | Sleep | Wake |
| **Mot.** | S | 1360 | 54 |
|  | W | 34 | 25 |

| (b) | Act. vs. Vivo. |
| --- | --- |
| **Sensitivity = $TP/(TP+FN)$** | 0.99 |
| **Specificity = $TN/(TN+FP)$** | 0.80 |
| **Accuracy = $(TP+TN)/(TP+TN+FP+FN)$** | 0.98 |

| Mot vs. Vivo. |
| --- |
| 1.00 |
| 0.83 |
| 0.99 |

| Act. vs. Mot. |
| --- |
| 0.98 |
| 0.32 |
| 0.94 |

**Table 3.** Parameter settings of the actigraphy report.

|  | **Proportional Integration Mode** | **Zero-Crossing Mode** |
| --- | --- | --- |
| **Epoch Length** | 0.5, or 1 min (default) | 0.5, or 1 min (default) |
| **Noise Threshold** | 0-100 mG (or default) | 0-100 mG (or default) |
| **Wake Threshold** | 20 (low), 40 (default), or 80 (high) | fixed |
| **Sleep Scoring Method** | Actiware algorithm [8] | Cole – Kripke algorithm [10] (default),<br>or Sadeh algorithm [11] |

### 3.2. Example Report

We offer a <u>sample actigraphy sleep report</u> for your reference [11]. This document outlines the parameter settings and provides a detailed analysis, as discussed in Section 3, including the step-by-step results.

Table 4 lists the parameter settings of the actigraphy report. Figure 7 shows the report of the one-night sleep profile and sleep parameters based on the default setting of zero-crossing mode listed in Table 4. Figure 8 shows the report of multiple participants with multiple nights.

**Figure 7.**
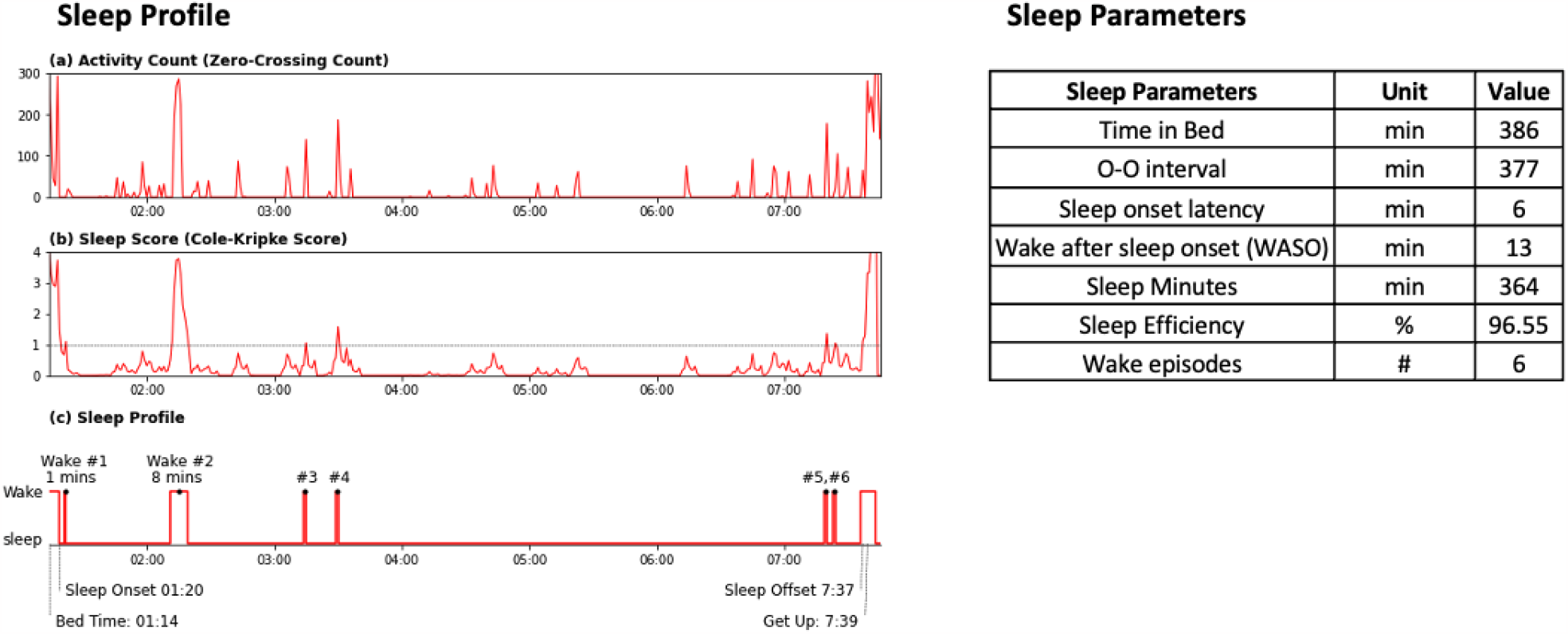
Report of the one-night sleep profile and sleep parameters.

**Figure 8.**
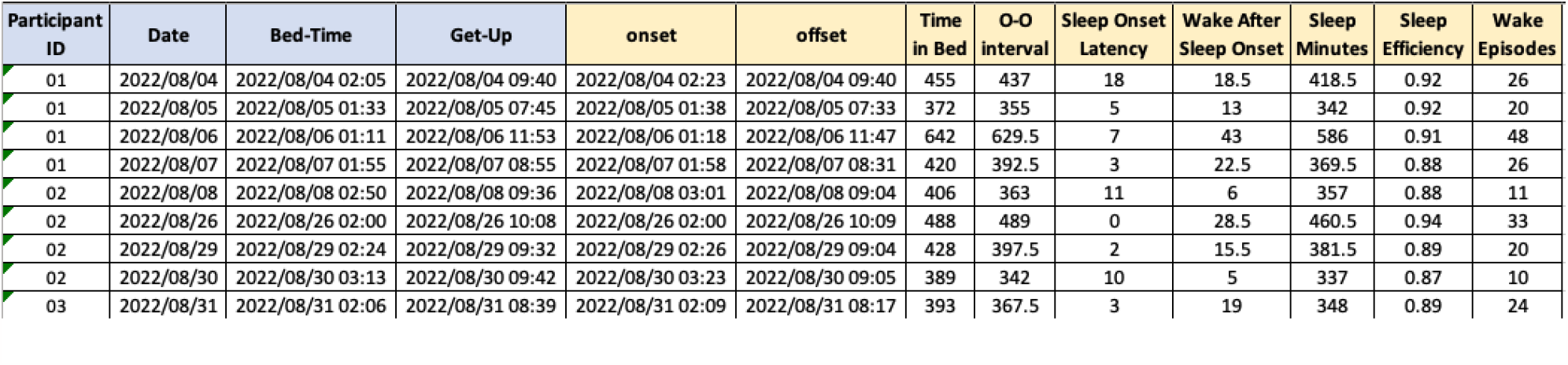
Report of multiple participants with multiple nights.

## 4. Conclusion

We have demonstrated that Garmin’s consumer-grade wearable devices can produce results closely aligned with those from medical-grade devices like Actiwatch-2 and Motionlogger. Specifically, when evaluated against these medical-grade devices, the Garmin device exhibited a sensitivity (for sleep detection), specificity (for wake detection), and overall accuracy of over 95%, 70%, and 95%, respectively. Additionally, we have detailed the transparent methodology used to generate sleep reports based on acceleration data and offer the flexibility for users to customize sleep/wake detection settings to meet personal needs.

Our findings also indicate a low level of consistency between Actiwatch-2 and Motionlogger, corroborating previous studies [12]. This inconsistency may arise from the complex interplay of different algorithms in Proportional Integrating Mode (PIM) and Zero Crossing Mode (ZCM). As noted by Fekedulegn et al., the choice of mode should consider various factors, such as age, gender, underlying medical conditions of the study population, and the specific research question at hand. As a result, decisions about mode selection and other parameter settings are likely to require customization and may vary significantly depending on the population studied [4].

## Data Availability

All data produced in the present study are available upon reasonable request to the authors.

## Acknowledgments

This work was supported by Labfront.

## Ethics approval

WIRB-Copernicus Group (WCG) approved the study protocol (number 20220365).

